# FKBPL and SIRT-1, key angiogenesis proteins, are downregulated by diabetes in pregnancy

**DOI:** 10.1101/2020.10.06.20208116

**Authors:** Abdelrahim Alqudah, Kelly-Ann Eastwood, Djurdja Jerotic, Naomi Todd, Denise Hoch, Ross McNally, Danilo Obradovic, Stefan Dugalic, Alyson J. Hunter, Valerie A. Holmes, David R. McCance, Ian S. Young, Chris J Watson, Tracy Robson, Gernot Desoye, David J Grieve, Lana McClements

## Abstract

**Context:** Diabetes in pregnancy is associated with numerous complications, however the mechanisms are still poorly understood.

**Objective:** To investigate the role of new angiogenesis markers, FKBPL and SIRT-1, in pre-gestational (type 1 diabetes, T1D) and gestational diabetes (GDM).

**Design and intervention:** Placental FKBPL, SIRT-1, PlGF and VEGF-R1 protein expression was determined from pregnant women with GDM or T1D, and in first trimester trophoblast cells exposed to high glucose and varying oxygen concentrations. Endothelial cell function was assessed in high glucose conditions and FKBPL overexpression.

**Settings and Participants:** Human placental samples from pregnant women with GDM (n=6) or T1D (n=8) were collected to assess FKBPL and SIRT-1 protein expression compared to non-diabetic controls.

**Main outcome measures:** To determine the role of placental FKBPL and/or SIRT-1 in diabetic pregnancies, in first trimester trophoblasts and endothelial cell function in high-glucose environment.

**Results:** Placental FKBPL protein expression was downregulated in T1D (FKBPL; p<0.05) whereas PlGF/VEGF-R1 were upregulated (p<0.05); correlations adjusted for gestational age were also significant. In the presence of GDM, only SIRT-1 (p<0.001) was significantly downregulated even when adjusted for gestational age (r=-0.92, p=0.001). FKBPL and SIRT-1 were also downregulated in ACH-3P cells in high glucose conditions and 6.5%/2.5% oxygen concentrations (p<0.05). FKBPL overexpression in HUVECs reduced tubule formation compared to empty vector control, in high glucose conditions (junctions; p<0.01, branches; p<0.05).

**Conclusions:** FKBPL and/or SIRT-1 downregulation in response to diabetes may have a role in the development of vascular dysfunction in pregnancy, and associated complications such as preeclampsia.

## Introduction

Hyperglycaemia is one of the most common pregnancy complications, affecting one in six pregnancies. ^1^ Hyperglycaemia in pregnancy includes new onset diabetes including gestational diabetes mellitus (GDM), and pre-existing diabetes in pregnancy (type 1 (T1D) or type 2 diabetes mellitus (T2D)), ^2^ with 86% of hyperglycaemia cases in pregnancy attributed to GDM. ^3^

Gestational hypertension and preeclampsia are the leading causes of morbidity and mortality in pregnancy, ^4^ and in the presence of hyperglycaemia, risk of these conditions is increased up to 4-fold. ^5^ Indeed, elevated fasting blood glucose and HbA1c have been associated with increased risk of preeclampsia in pregnant women with GDM, T1D or T2D. ^6–8^ In addition, hyperglycaemia during pregnancy can lead to several other adverse neonatal complications, including the large for gestational age (LGA) foetus, shoulder dystocia, respiratory distress and neonatal hypoglycaemia. ^2^ Hyperglycaemia in pregnancy also increases the risk of perinatal death and stillbirth by at least two-to three-fold, particularly in the presence of T1D and T2D compared to healthy pregnant controls.^9,10^ Women who develop GDM in pregnancy have a 50% increased risk of developing GDM in a subsequent pregnancy, and a 10-fold increased risk of developing T2D within 10 years following pregnancy. ^11^ In addition, children born to mothers with GDM or pre-existing diabetes are at higher risk of developing obesity, T2D and cardiovascular disease during their life-time. ^12–14^ Despite clear epidemiological link between diabetes in pregnancy and both short- and long-term pregnancy complications, the mechanisms of these associations are still poorly understood.

The placenta is a highly vascularised organ, which facilitates adequate oxygen and nutrient transfer from the mother to foetus as well as removal of foetal waste products through maternal circulation. ^15^ Placental development is tightly regulated by angiogenic factors, such as vascular endothelial growth factor (VEGF), placental growth factor (PlGF), and their receptors, which stimulate angiogenesis throughout pregnancy in a controlled manner. ^16^ However, in pregnancies complicated by diabetes, pre-existing endothelial dysfunction and aberrant angiogenesis can lead to hyper-vascularisation of the placenta. ^17^ This process of overstimulated placental angiogenesis can lead to impairment in the integrity of both the maternal and foetal vascular system as well as increased peripheral vascular resistance and maternal hypertension. ^18^

FK506-binding protein like (FKBPL), a novel member of the immunophilin protein family, is a key secreted anti-angiogenic protein that also regulates glucocorticoid receptor signalling. ^19–22^ Both of these key functions regulate metabolism and vascular health, ^23,24^ therefore suggesting a potentially important role for FKBPL in diabetes and the associated vascular dysfunction. Sirtuin-1 (SIRT-1), an NAD+ dependant deacetylase, also has a critical role in endothelial cell metabolism and function. In diabetes, SIRT-1 is downregulated, which is often associated with endothelial dysfunction. ^25,26^ Previous studies have also demonstrated that FKBPL knockout mice were embryonically lethal, highlighting its vital role in embryonic development. Whilst heterozygous FKBPL knockdown mice developed normally, the vasculature appeared leaky with compromised integrity, implicating FKBPL as a key regulator of endothelial function. ^27^ A FKBPL has also been demonstrated a key role in tumour angiogenesis; this has led to the development of therapeutic peptides for the treatment of solid tumours. ^28,29^ In preeclampsia, FKBPL and its target protein, CD44, have demonstrated their predictive and diagnostic biomarker potential, reflective of the pathogenesis of preeclampsia and placental oxygen changes throughout gestation. ^30,31^ However, FKBPL’s role in the context of diabetes in pregnancy has not yet been investigated. Therefore, in this study, placental expression of FKBPL in conjunction with SIRT-1, PlGF and VEGFR1 was investigated in human samples from pregnant women with pre-existing diabetes (T1D), and GDM. FKBPL expression was also assessed in ACH-3P first trimester trophoblasts in response to high glucose and varying oxygen concentrations relevant to placental development. ^31^

## Methods

### Placental samples

Pregnant women with GDM and T1D as well as healthy pregnant women, provided written informed consent as part of their recruitment to the PREDICT study at the Royal Jubilee Maternity Hospital ^32^. Ethical approval for this project was obtained from the NHS Health Research Authority (ORECNI, 14/NW/1222) and the School of Medicine, Dentistry and Biomedical Sciences (Queen’s University Belfast). Clinical characteristics were recorded (age, BMI and gestational age (GA) and foetal sex) and systolic blood pressure (sBP) and diastolic blood pressure (dBP) was measured according to the National Heart Foundation of Australia protocol using automated devices as previously described ^33^ in both the left and right arm of each patient on two occasions during the first trimester. Placental sections were collected from women with T1D, GDM or non-diabetic pregnancies after the baby was delivered.

All slides were stained by standard haematoxylin and eosin (H&E) procedure. Following staining, placental morphology and pathophysiology was assessed by a pathologist, DO. The parameters analysed included placental maturity, vascularisation and branching of chorionic villi by characterising the structure of chorionic villi, area of terminal villi covered by blood vessels, presence or absence of calcifications and syncytial knots. The distribution of syncytial knots was quantified by counting the number of syncytial knots per 100 terminal villi.

### Immunofluorescence staining

Placental section slides were prepared by the Northern Ireland Biobank ^34^ using fresh tissue before being subjected to immunofluorescence staining for FKBPL (Cat. no.: 10060-1-AP; Proteintech, UK), SIRT-1 (Cat. no.: Ab110304; Abcam, UK), PlGF (Cat. no.: ab180734, Abcam, UK) and VEGFR1 (Cat. no.: AF321, R&D systems, USA). Tissue slides were imaged using a Leica DMi8 fluorescence microscope using the same magnification (20x) and exposure. Analysis was performed using image J software (NIH, US) by selecting six random fields of view per section and measuring the intensity, with the assessor blind to patient group. Protein expression was quantified as previously described. ^35^

### Cell culture

ACH-3P cells were kindly donated by Professor Gernot Desoye (Graz Medical University, Austria). Briefly, immortalised choriocarcinoma cells, AC1-1 cells, and primary trophoblast cells isolated from the first trimester placenta were fused^36^, to form a unique ACH-3P cell line as previously described^37^. ACH-3P cells were maintained in Hams F-12 medium supplemented with 10% FBS and all experiments were carried out at Graz Medical University. Short Tandem Repeat (STR) DNA profiling analysis was performed using PowerPlex 16 HS System (Promega, UK) for cell authentication. ACH-3P cells were exposed to high glucose (25 mM) or mannitol (20 mM + 5 mM D-glucose) as an osmolality control under different oxygen concentrations (21%, 6.5%, and 2.5%) for 24 h. Protein was then extracted for downstream protein expression analysis by Western blotting. All experiments with ACH-3P cells were carried out at the Graz Medical University.

Human Umbilical Vein Endothelial Cells (HUVECs) (ATCC, USA) were kindly donated by Dr Andriana Margariti (Queen’s University Belfast) were maintained in MV2 endothelial cell growth media (PromoCell, Germany) supplemented with low serum growth supplement containing the following: 5% foetal calf serum (FCS), epidermal growth factor 5 ng/ml, basic fibroblast growth factor (FGF) 10 ng/ml, insulin-like growth factor 20 ng/ml, VEGF 0.5 ng/ml, ascorbic acid 1 μg/ml and hydrocortisone 0.2 μg/ml. All cells were maintained at 37 °C in a humidified atmosphere with 5 % CO_2_.

### Tubule Formation Assay

HUVECs were treated with a FKBPL overexpressing plasmid (pFKBPL; Sino Biological, USA) or empty vector plasmid (pCMV3; Sino Biological, USA) for 24 h before being plated in Matrigel. HUVEC (7×10^5^) were stained with calcein (2µg/ml; Thermo Fisher Scientific, UK) prior to being seeded on phenol red free reduced-growth factor Matrigel under high glucose conditions (30mM D-glucose) or mannitol (25 mM + 5 mM D-glucose) for 24 h. Tubule formation was imaged by randomly capturing 6 images per well using a DMI8 inverted florescence microscope. The number of branches and junctions were quantified using Image J software (NIH, USA).

### Western blotting

HUVEC and ACH-3P cell lysates were harvested using RIPA buffer (SantaCruz Biotechnology, USA) supplemented with protease and phosphatase inhibitor cocktails (Roche, UK) and subjected to western blotting. FKBPL (Protientech, UK, cat# 10060-1-AP) and SIRT-1 (Abcam, UK, cat# Ab110304) antibodies were used at 1:1000 dilution.

### Statistical Analysis

Analysis of normally distributed data was undertaken using two-tailed unpaired t-tests. For skewed data, Mann Whitney comparisons test were performed. Results are expressed as the mean ± the standard error of the mean (SEM) and values were considered statistically significant if p<0.05. Analyses were performed using Prism 5 software (GraphPad Software, La Jolla, CA, USA). Pearson correlation analysis was performed to test the correlation between the presence of diabetes and placental protein expression adjusted for gestational age using SPSS 20.0 (IBM, USA).

## Results

### Signs of placental hypo-perfusion and increased vascularisation are evident in diabetes

In order to assess placental morphology in pregnant women with diabetes, H&E staining was performed on placental sections and quantification of syncytial knots performed. Increased numbers of syncytial knots, aggregates of syncytial nuclei at the surface of terminal villi^38^, can be indicative of the presence of placental hypoxia. However, advancing gestation is also consistent with an increased presence of syncytial knots. These structures can therefore be used to evaluate villous maturity. ^39^ In normal placentae, approximately 30% of terminal villi have syncytial knots, whilst a higher percentage of syncytial knots is associated with uteroplacental malperfusion. ^40^ In twelve out of fourteen placental samples from pregnant women with diabetes (T1D and GDM), an increased number of syncytial knots was observed compared to normal pregnancies [Figure 1 (A; T1D, n=8, p<0.05), B; GDM, n=6, p<0.05)]. Placental syncytial knots were positively correlated with T1D (r=0.619, p=0.024), which remained significant when adjusted for gestational age (r=0.66, p=0.02). Although syncytial knots were positively correlated with GDM (r=0.781, p=0.013), this correlation became non-significant when correcting to gestational age (r=0.459, p=0.252). Moreover, in all fourteen placental samples from diabetic pregnancies, a general increase in villous vascularity was observed, often associated with villous immaturity. This suggests that there is an increased number of capillaries and macrophages with the presence of fluid within the villous structure, which is a well-recognised feature of maternal diabetes (Figure 1C). ^41^ Furthermore, overstimulated placental angiogenesis can lead to impairment of the integrity of the vascular system and increased resistance to blood flow leading to hypoxia and the observed syncytial knots; consistent with the observed increased pro-angiogenic and reduced anti-angiogenic factors in these tissues. ^18^ No differences in calcification were observed between diabetic and non-diabetic placentae (Figure 1C).

**Figure 1.**
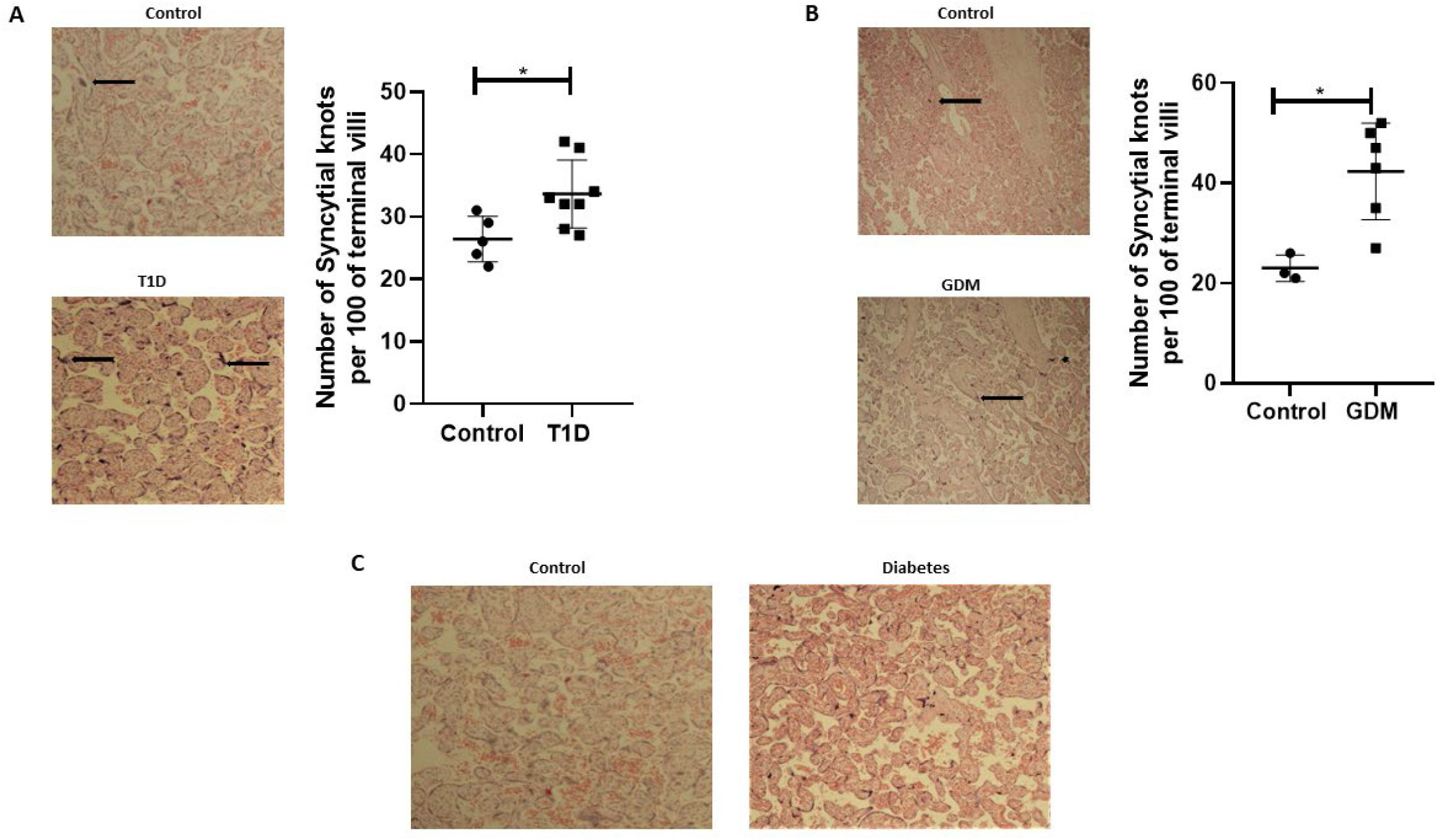
Sign of placenta hypoxia is evident concomitantly with increased levels of villous vascularity in diabetes. Paraffin-embedded placental sections were stained with H&E with 2 separate fields/section imaged at 4x magnification. **(A&B)** The number of syncytial knots which are aggregates of syncytial nuclei at the surface of terminal villi (black spots indicated by arrows) is increased in placental samples from women with T1D or GDM compared to non-diabetic controls (T1D, n=8, GDM, n=6 two tailed paired t-test, *<0.05). **(C)** Villous vascularity (indicated by increased red staining) is abundant in diabetes compared to non-diabetic controls.

### Placental FKBPL and SIRT-1 expression is reduced in diabetic pregnancies

Placental samples were collected from 14 pregnant women, six of whom developed GDM and eight of whom had pre-existing T1D. Control placental samples (n=8) were collected from healthy pregnant women matched for BMI and age, 1:1. Baseline characteristics are shown in Table 1. No statistically significant differences in age, BMI, blood pressure or foetal sex were noted between any of the study groups compared to their matched controls. However, gestational age at delivery was lower in T1D and GDM patients compared to control.

**Table 1.** Maternal baseline characteristics for pregnant women with type 1 diabetes and gestational diabetes.

|  | **T1D (n=8)** | **Control (n=5)** | **p value** |
| --- | --- | --- | --- |
| BMI | 28.0±6.9 | 25.9±5.7 | 0.58 |
| Age | 24.6±5.7 | 28.8±4.8 | 0.20 |
| sBP | 115±8.8 | 114±8.7 | 0.92 |
| dBP | 73.8±9.1 | 69.8±24.4 | 0.36 |
| Gestational age<br>(weeks) | 35.38±0.8 | 39±1.4 | 0.009 |
| Foetal sex | 2 females<br>6 males | 1 female<br>4 males | 0.85 |
|  | **GDM (n=6)** | **Control (n=3)** | **p value** |
| BMI | 32.7±8.9 | 29.34±4.4 | 0.29 |
| Age | 31.8±3.8 | 27±2.6 | 0.06 |
| sBP | 122.1±8.1 | 121.6±1.8 | 0.9 |
| dBP | 85.3±6.8 | 78.3±8.1 | 0.16 |
| Gestational age<br>(weeks) | 36.8±1.2 | 39±0 | 0.02 |
| Foetal sex | 2 females<br>4 males | 3 males | 0.17 |

Data is presented as mean ± SD, two tailed paired Student's t-test. *Foetal sex was calculated by assuming female is zero and male is one. BMI-Body mass index; sBP-Systolic blood pressure; dBP-Diastolic blood pressure*

To assess the role of FKBPL and SIRT-1 in pregnancies complicated with diabetes, protein expression of FKBPL and SIRT-1 within placental samples collected from pregnant women with T1D or GDM was compared to controls, matched for age and BMI. Our results demonstrated significant downregulation of FKBPL protein expression within placental sections of T1D patients compared to non-diabetic controls (Figure 2A, n=8, p<0.05). FKBPL placental expression was negatively correlated with T1D (Table 2, r=-0.581, p=0.037), which remained significant when correcting for gestational age (Table 2, r=-0.65, p=0.022). SIRT-1 protein expression remained the same within the placental sections in the presence of T1D compared to controls (Figure 2B, n=8). No significant correlation was observed between placental SIRT-1 expression and T1D (Table 2, r=-0.428, p=0.145), however this correlation became borderline significant when adjusted for gestational age (r=-0.578, p=0.049).

**Table 2.**
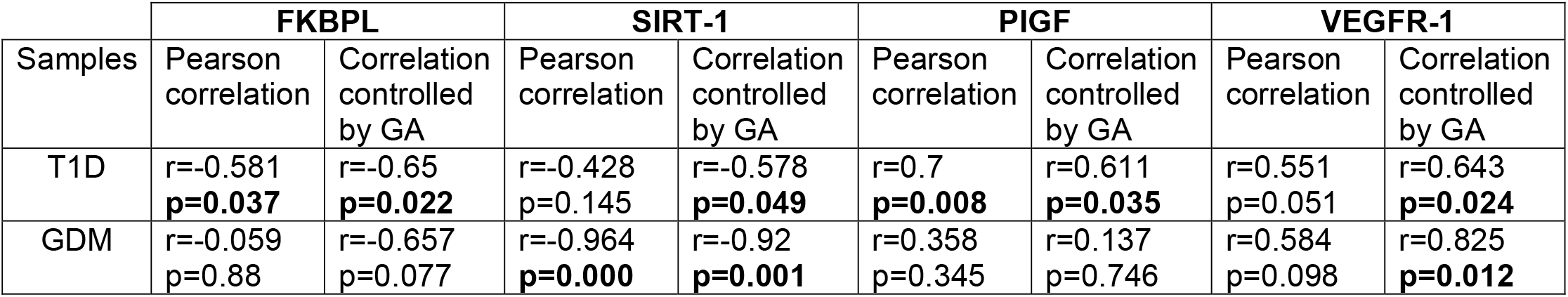
Adjusted correlations for differences in gestational age between diabetic and non-diabetic placentae.

| Samples | FKBPL |  | SIRT-1 |  | PIGF |  | VEGFR-1 |  |
| --- | --- | --- | --- | --- | --- | --- | --- | --- |
|  | Pearson correlation | Correlation controlled by GA | Pearson correlation | Correlation controlled by GA | Pearson correlation | Correlation controlled by GA | Pearson correlation | Correlation controlled by GA |
| T1D | r=-0.581<br>**p=0.037** | r=-0.65<br>**p=0.022** | r=-0.428<br>p=0.145 | r=-0.578<br>**p=0.049** | r=0.7<br>**p=0.008** | r=0.611<br>**p=0.035** | r=0.551<br>p=0.051 | r=0.643<br>**p=0.024** |
| GDM | r=-0.059<br>p=0.88 | r=-0.657<br>p=0.077 | r=-0.964<br>**p=0.000** | r=-0.92<br>**p=0.001** | r=0.358<br>p=0.345 | r=0.137<br>p=0.746 | r=0.584<br>p=0.098 | r=0.825<br>**p=0.012** |

**Figure 2.**
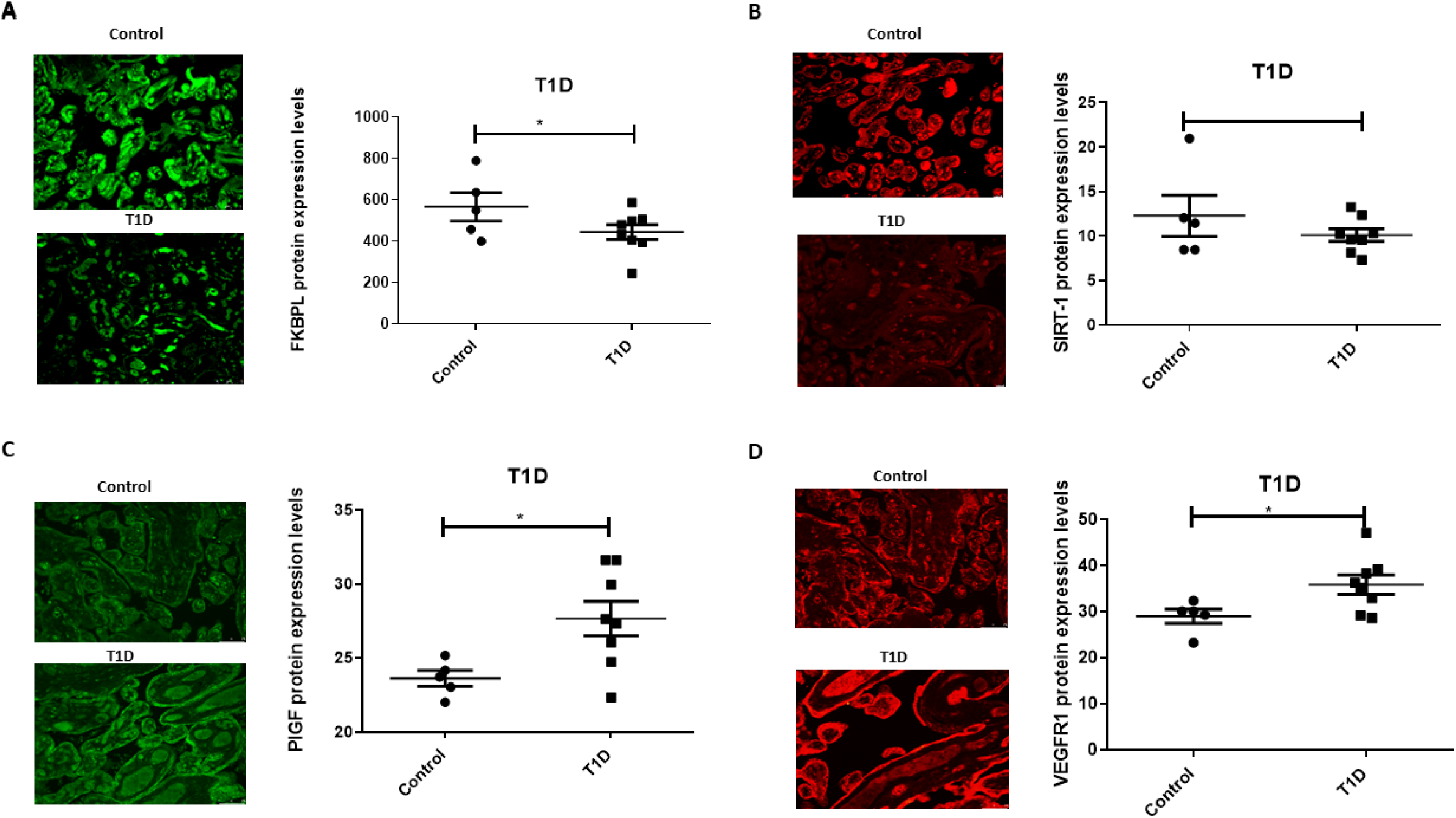
Placental angiogenic balance is disrupted in T1D. Placental slides from women with T1D versus age, BMI and foetal sex matched controls (one-to-one match) were stained with **(A)** FKBPL, **(B)** SIRT-1, **(C)** PlGF and **(D)** VEGFR1 primary antibodies, followed by staining with green Alexaflour or red Cy3 secondary antibody. Six images per slide were taken at 20x magnification and the mean fluorescence quantified using Image J. Representative images inset. FKBPL/PlGF: two tailed unpaired t-test, n=8; VEGFR1: Mann-Whitney, *<0.05).

Interestingly, PlGF protein expression was upregulated in placentae collected from women with pre-existing T1D (Figure 2C, n=8, p<0.05) with a similar pattern observed for VEGFR-1 protein (Figure 2D, n=8, p<0.05). PlGF expression in the placenta was positively correlated with T1D (Table 2, r=0.7, p=0.008), which remained significant when correcting for gestational age (Table 2, r=0.611, p=0.035). Similarly, VEGFR-1 expression was positively correlated with T1D with borderline significance (Table 2, r=0.551, p=0.051), however after adjusting for gestational age, this correlation was statistically significant (Table 2, r=0.643, p=0.024).

In the presence of GDM, placental FKBPL protein expression remained unchanged compared to non-diabetic controls (Figure 3A, n=6) with non-significant negative correlation (Table 2, r=-0.059, p=0.88), without further improvement after correcting for gestational age (Table 2, r=-0.657, p=0.077). GDM led to reduced SIRT-1 protein expression in the placenta (Figure 3B, n=6, p<0.001). SIRT-1 expression was negatively correlated with GDM (Table 2, r=-0.964, p=0.000), which remained significant after adjusting for gestational age (Table 2, r=-0.92, p=0.001). However, in contrast to T1D and similar to FKBPL, placental PlGF and VEGFR-1 protein expression remained unchanged in the presence of GDM compared to controls (Figure 3C&D, n=6). There was no significant correlation between PlGF expression and GDM (Table 2, r=0.358, p=0.345), which remained non-significant after correcting the values to gestational diabetes (Table 2, r=0.137, p=0.746). Similarly, the correlation between VEGFR-1 expression and GDM within the placental sections was not significant (Table 2, r=0.584, p=0.098) however, there was a significant positive correlation once adjusted for gestational age (Table 2, r=0.825, p=0.012).

**Figure 3.**
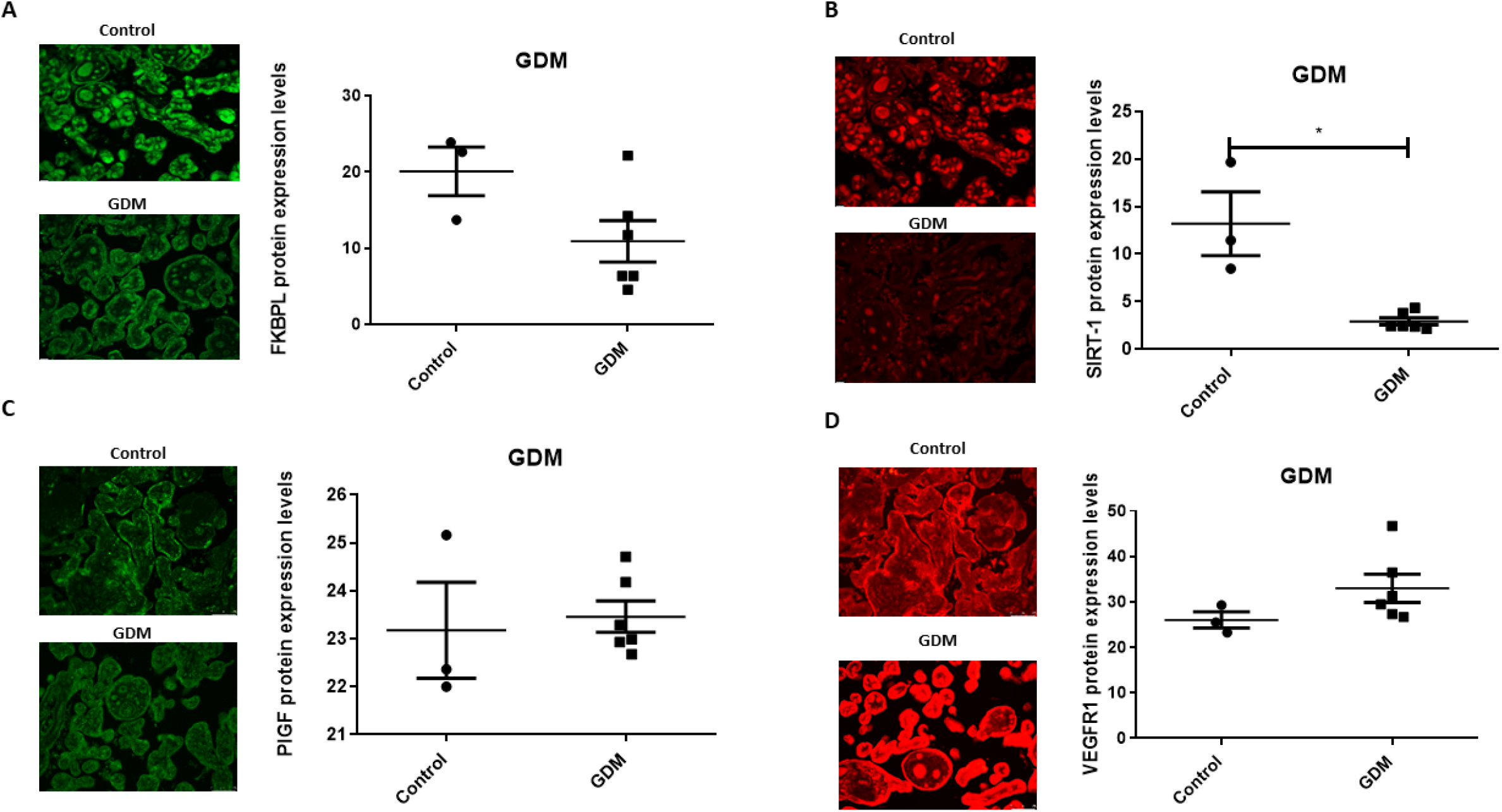
Placental angiogenic balance is distupted in GDM. Placental slides from women with GDM versus age, BMI and foetal sex matched controls (one-to-one match) were stained with **(A)** FKBPL, **(B)** SIRT-1, **(C)** PlGF and **(D)** VEGFR1 primary antibodies, followed by staining with green Alexaflour or red Cy3 secondary antibody. Six images per slide were taken at 20x magnification and the mean fluorescence quantified using Image J. Representative images inset. SIRT-1: Mann-Whitney test; n=6, *<0.001.

### FKBPL and SIRT-1 protein expression are downregulated in response to high glucose and low oxygen in ACH-3P cells

Having demonstrated that FKBPL and SIRT-1 are downregulated in the presence of T1D and GDM within the placentae collected following delivery, we wanted to investigate their regulation by diabetic environment early in pregnancy. For this purpose, we used a custom-made first-trimester trophoblast cell line, ACH-3P, generated by fusing primary first trimester trophoblasts with the choriocarcinoma cell line, AC1-1, so thereby closely resembling primary extravillous trophoblasts. ^36,37^ In order to mimic conditions associated with early pregnancy and placental development, ACH-3P cells were first exposed to varying oxygen (O_2_) concentrations (21%, 6.5% and 2.5%), similar to oxygen tension observed in the first trimester during placental development (6.5%), and 2.5% oxygen, which represent hypoxia conditions ^42–44^. To investigate the effect of a diabetic environment on FKBPL and SIRT-1 protein expression, ACH-3P cells were also exposed to high glucose (25 mM D-glucose) or normal glucose concentration (5 mM D-glucose + 20 mM Mannitol) in the presence of varying oxygen levels (21%, 6.5% and 2.5%) to mimic hyperglycaemic conditions in the first trimester of diabetic pregnancy. The effect of varying oxygen concentrations in ACH-3P cells was observed between 21% and 2.5% O_2_ in relation to FKBPL and SIRT-1 protein expression (Figure 4A, n=3, p<0.05), and between 6.5% and 2.5% O_2_ in relation to SIRT-1 protein expression (Figure 4A, n=3, p<0.05). FKBPL protein expression was not changed when ACH-3P cells were exposed to high glucose under high oxygen conditions (21% O_2_) (Figure 4B, n=3). Similarly, SIRT-1 protein expression also remained unchanged when ACH-3P cells were exposed to the same hyperglycaemic and normoxic conditions (Figure 4B, n=3). However, under low oxygen tensions of 6.5% (Figure 4C) and 2.5% O_2_ (Figure 4D), both FKBPL (n=3, p<0.05) and SIRT-1 (n=3, p<0.05) were downregulated, suggesting that hypoxia can decrease the levels of these proteins, consistent with hypoxia promoting a pro-angiogenic phenotype.

**Figure 4.**
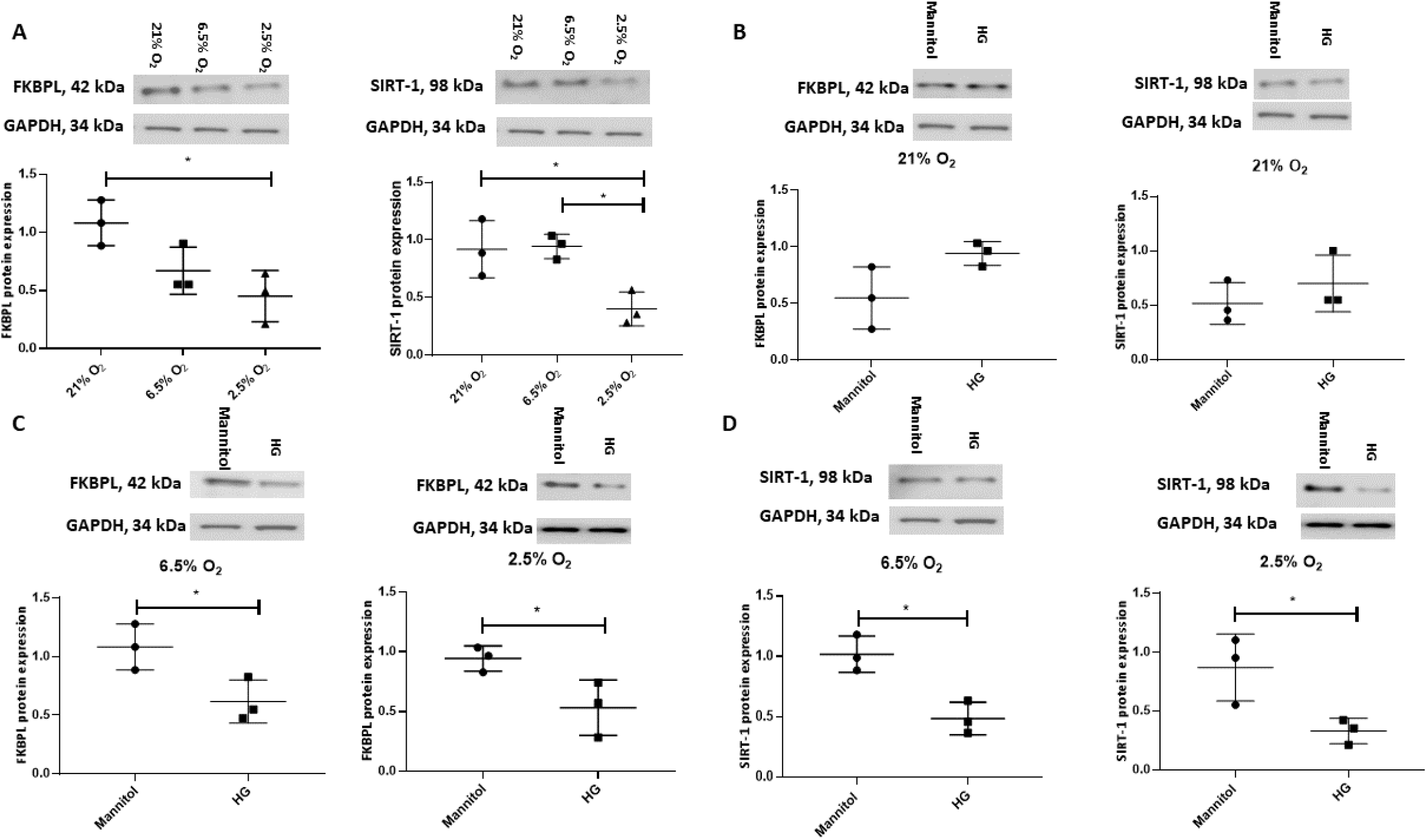
FKBPL and SIRT-1 protein expression is reduced in ACH-3P cells in high glucose conditions with low oxygen levels. ACH-3P cells were exposed to high glucose (25mM) in the presence of varying O_2_ conditions for 24 h before protein was extracted and western blotting performed. **(A)** FKBPL and SIRT-1 protein expression is reduced in low oxygen (2.5%) conditions at normal glucose levels. **(B)** FKBPL and SIRT-1 protein expression remained unchanged in high oxygen conditions in the presence of normal glucose. **(C)** FKBPL and **(D)** SIRT-1 protein expression was reduced in high glucose conditions at 6.5% and 2.5% O_2_ compared to normal glucose conditions at the same oxygen levels. Protein expression was normalised to GAPDH. (n=3, A&B one-way ANOVA with Bonferroni post-hoc test’ C&D unpaired Student’s t-test, *<0.05).

### FKBPL plays a key role in regulating endothelial cell angiogenic potential in hyperglycaemia

To determine whether overexpression of FKBPL in hyperglycaemic conditions would restore normal angiogenesis in diabetic pregnancies, HUVECs were treated with a FKBPL overexpressing plasmid (pFKBPL) or empty vector control plasmid (pCMV3) for 24 h before being plated in Matrigel in the presence of high glucose media (30 mM) or normal glucose media (5.5 mM + 24.5 mM mannitol) for 24 h. The number of junctions and branches formed was lower with FKBPL overexpression, consistent with its anti-angiogenic properties, in normal glucose conditions compared to empty vector control [Figure 7, n=6, p<0.01 (junctions), p<0.001 (branches)]. High glucose environment itself led to a similar reduction in endothelial cell angiogenic potential [Figure 7B, n=6, p<0.01 (junctions), p<0.001 (branches)]. Furthermore, when FKBPL was overexpressed in high glucose conditions, the number of junctions and branches was further reduced compared to empty vector high glucose control (Figure 5A, n=6, p<0.01 (junctions), p<0.05 (branches)]. In addition, the number of junctions and branches was lower with FKBPL overexpression in high glucose media compared to FKBPL overexpression in normal glucose media (Figure 5B, n=6, p<0.01). This was performed at experimental oxygen conditions of 21%. High glucose therefore appears to potentiate the anti-angiogenic action of FKBPL.

**Figure 5.**
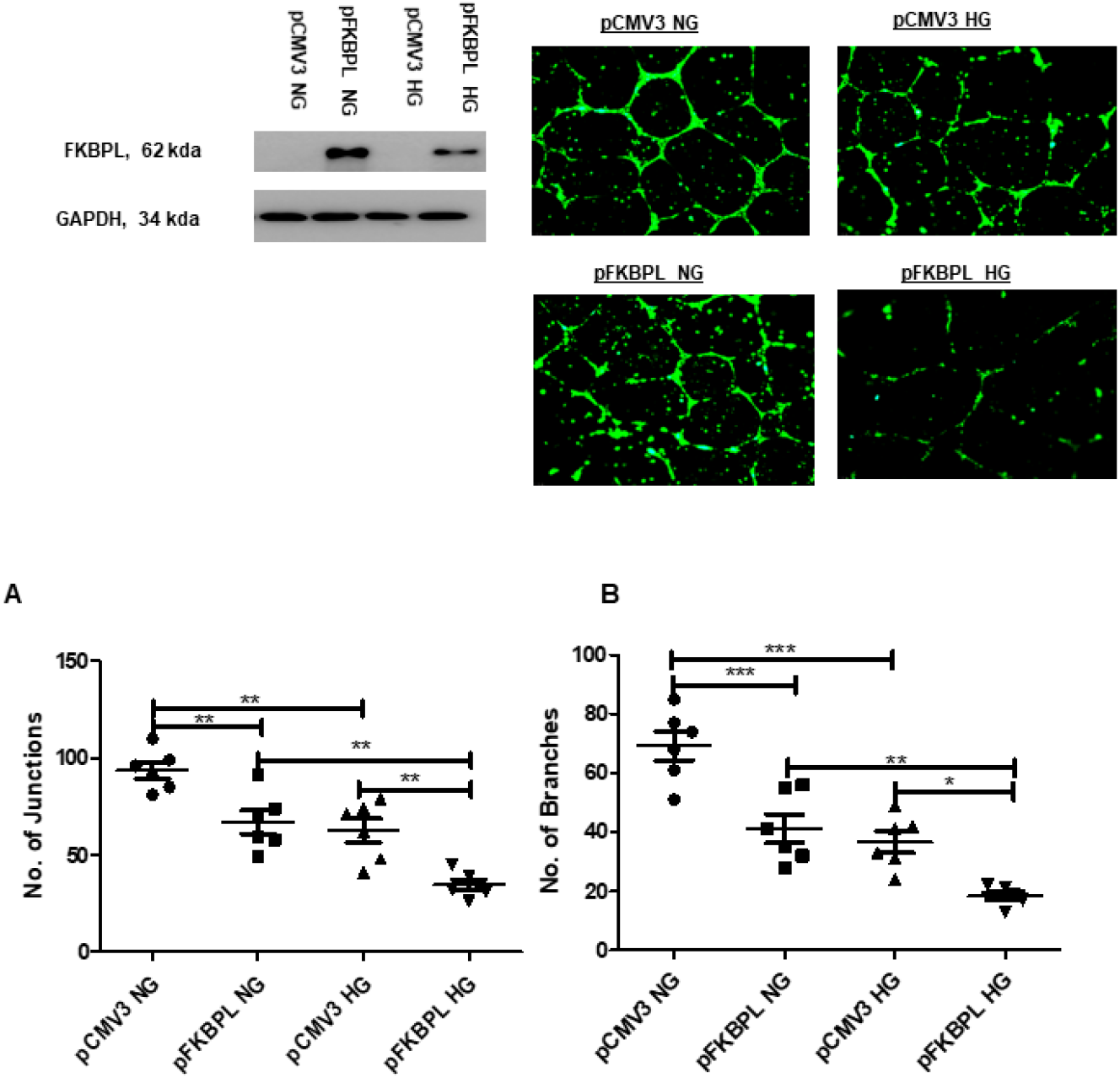
Angiogenic potential is reduced with FKBPL overexpression in high glucose conditions in HUVECs. The number of **(A)** junctions and **(B)** branches was quantified following FKBPL overexpression in HUVECs for 24 h. The cells were stained with calcein stain, before being plated in Matrigel and exposed to high glucose (HG, 30 mM) or normal glucose (NG, 5.5 mM) media for 24 h. Six images/well were taken using a DMi8 microscope; representative images are shown inset. Images were analysed based on the number of junctions and branches formed using ImageJ angiogenesis macros. (n=6, *<0.05, **<0.01, ***<0.001, One-way ANOVA, Bonferroni multiple comparison test).

## Discussion

In this study, we demonstrated that expression of the key proteins in placental angiogenesis, FKBPL, SIRT-1, PlGF and VEGFR-1, ^27,30,45–47^ is dysregulated in pregnancies complicated by diabetes. The FKBPL’s role in diabetic pregnancies has never been demonstrated previously. Thus, our study demonstrates, for the first time, that FKBPL, a key anti-angiogenic protein, is downregulated in T1D. We also show that PlGF and VEGFR-1 are upregulated in T1D, which represents an aberrant but overstimulated angiogenesis profile, also supported by placental histology results from diabetic pregnancies. No changes in these proteins’ expression were observed in GDM cohort, which could be due to a low number of samples in this group (n=6, GDM and n=3, non-diabetic controls) given that there was a trend observed with FKBPL and VEGFR-1.

While SIRT-1 expression was not affected by T1D, it was significantly downregulated in the presence of GDM. The role of SIRT-1 in trophoblast function, important for placental development has been demonstrated before. ^48^ Our work also validates the previous findings that SIRT-1 is a key pathway affected by GDM. ^49^

Furthermore, we demonstrated in relevant *in vitro* models that FKBPL and SIRT-1 proteins are reduced in both the presence of hyperglycaemia and low oxygen tension that could have adverse effects on trophoblast function and placental development^31^. FKBPL showed a critical role in endothelial angiogenic potential in both normal and high glucose environment. This is important as the placenta is a highly vascularised organ with a crucial role in supporting growth and development of the foetus. ^15^ The placental circulation is divided into two separate systems: 1) the maternal-placental circulation where the maternal blood flows through spiral uterine arteries to the intervillous space surrounded by the terminal villi to facilitate oxygen and nutrient exchange, and 2) the foetal-placental circulation, which allows the umbilical arteries to carry de-oxygenated blood from the foetus to the villous vessels. ^50^ Therefore, controlled angiogenesis is key for normal placental development during pregnancy and any aberrant changes in angiogenic balance are closely associated with development of pregnancy complications such as preeclampsia. Dysregulation in FKBPL, SIRT-1, PlGF and VEGFR-1 demonstrated in this study, albeit with differential regulation in T1D or GDM, indicate the presence of angiogenic imbalance in pregnancies complicated by diabetes. Further studies should investigate how this affects trophoblast function and placental development.

Our previous study using FKBPL knockdown mice demonstrated strong pro-angiogenic phenotype with early signs of endothelial dysfunction. ^27^ Findings from this study support an anti-angiogenic role of FKBPL in endothelial cells, however our data also suggest that low levels of endothelial FKBPL could be beneficial in diabetic pregnancies. This is because high levels of FKBPL appeared to cause further restriction of angiogenesis in hyperglycaemic conditions. Further investigations need to be conducted to determine the mechanisms and effects of FKBPL on the integrity of endothelial barrier. Moreover, SIRT-1, an important mediator of endothelial function, has been previously reported to be decreased in diabetes and associated with endothelial dysfunction, ^51^ further supporting the presence of endothelial dysfunction in pregnancies complicated by diabetes potentially due to reduced levels of SIRT-1 and FKBPL. Indeed, endothelial dysfunction has been previously associated with diabetes and other related complications ^52,53^.

It is well-established that maternal diabetes affects placental vascular development and that is dependent upon the duration and type of diabetes. ^54^ Pregnant women with T1D may affect the entire period of placental and foetal development, whereas GDM may only impact placental growth in the later stages of pregnancy. ^55^ Diabetic placentation is characterised by increased angiogenesis due to raised VEGF levels. This leads to alterations in the integrity of the maternal and foetal vascular system, resulting in increased vascular permeability and vascular resistance ^18,56^. Our findings indicate that in diabetic placentae, there is an increase in syncytial knots indicative of hypoxia, in association with increased placental vascularity and immaturity. This could potentially be linked to increased PlGF and VEGF or decreased FKBPL or SIRT-1 expression as demonstrated in our study, promoting development of immature and leaky capillaries in the placenta. ^57^ Considering that FKBPL was downregulated in T1D placentae, and that angiogenic potential was further reduced with FKBPL overexpression in high glucose conditions, this may suggest its important role in regulating placental angiogenesis.

Trophoblast migration and invasion represent key processes driving placental development, particularly given remodelling of spiral uterine in pregnancy. ^58^ We have previously shown that the FKBPL plasma levels are reduced before the onset of preeclampsia and increased following the diagnosis of preeclampsia. ^30^ In this study, we demonstrated that FKBPL expression in ACH-3P first trimester trophoblast cells is downregulated by varying oxygen levels, recapitulating both normal and hypoxic conditions of the first trimester during placental development^31^. The observed reduction in FKBPL levels could therefore play a role in promoting migration and invasion of trophoblast cells. However, we have previously shown that overexpression of FKBPL inhibited migration and invasion of cancer cells. ^21,59^ Considering that hyperglycaemia has the potential to decrease trophoblast invasion, ^60^ FKBPL expression was found to be reduced in high glucose conditions, but only in the presence of low oxygen levels, which suggests that FKBPL may be important in determining primary trophoblast invasion during the first trimester of pregnancy in women with diabetes.

The limitations of the study include the small number of placentae in diabetes and non-diabetic groups and differences in gestational age between the groups. Nevertheless, using adjusted correlations we accounted for these limitations. We also demonstrated similar findings in terms of FKBPL and SIRT-1 regulation using our *in vitro* models of first trimester trophoblast and endothelial cell assay in high glucose environment representative of diabetic pregnancies.

In conclusion, placental FKBPL and SIRT-1 expression appears to be downregulated in response to diabetes in T1D and GDM, respectively, as well as following exposure to high glucose in trophoblast cells only in low oxygen conditions. This might suggest that carefully restoring FKBPL and SIRT-1 to normal physiological levels in diabetic conditions may promote trophoblast invasion and reduce hyper-vascularisation, with a potential to prevent subsequent pregnancy complications such as preeclampsia associated with diabetes.

## Data Availability

The datasets generated and/or analyzed during the current study are not publicly available but are available from the corresponding author upon reasonable request.

## Acknowledgements

We thank Dr Andriana Margariti for kindly donating human umbilical cord endothelial cells or HUVECs for cell function studies. We thank the PREDICT study participants and team for collection and provision of placental samples. We thank Northern Ireland Biobank for facilitating processing of the PREDICT placental samples.

